# Modelling the impact of combining HIV prevention interventions on HIV dynamics in fishing communities in Uganda

**DOI:** 10.1101/2022.05.20.22275218

**Authors:** Cécile Kremer, Anatoli Kamali, Monica Kuteesa, Janet Seeley, Niel Hens, Rebecca N Nsubuga

## Abstract

**Background:** In countries with mature generalized HIV epidemics such as Uganda, there are still groups of individuals that are disproportionately affected. These most at risk populations (MARPs) have high HIV prevalence compared to the general population. Among the MARPs in Uganda are fishing communities, which make up about 10% of the population.

**Methods:** We investigated the impact of combined HIV prevention interventions on HIV transmission dynamics in high-risk fishing communities in Uganda using a deterministic compartmental model. The model was calibrated to seroprevalence data from a census performed in 2014. To account for remaining uncertainty in the calibrated model parameters, an additional 50 000 simulated scenarios were modelled to investigate the impact of combined interventions.

**Results:** The projected HIV incidence decreased from 1.87 per 100 PY without intervention scale-up to 0.25 per 100 PY after 15 years (2014-2029) of intervention scale-up. A potential combination achieving this 87% reduction in incidence included condom use in about 60% of sexual acts, 23% of susceptible men circumcised, 87% of people living with HIV aware of their status, 75% of those on ART, and about 3% of susceptible individuals on oral PrEP. Uncertainty analysis revealed relative reductions in incidence ranging from 30.9% to 86.8%. Sensitivity analyses suggested that condom use and early ART were the most important intervention components.

**Conclusions:** Reducing HIV incidence, and consequently prevalence and AIDS-related mortality, in these high-risk fishing communities in Uganda is attainable in the long-run with a combination prevention package. Our projected intervention coverage levels are well within the national targets set by the Uganda government and close to reaching the UNAIDS 95-95-95 goal to end AIDS by 2030.

## Background

With 1.5 million new infections in 2021, HIV-1 remains a major global health problem. Of these new infections, 45% occurred in Eastern and Southern Africa (1). In countries with mature generalized epidemics such as Uganda, there are still groups of individuals that are disproportionately affected by HIV. These most at risk populations (MARPs) have high HIV prevalence compared to the general population, where prevalence has remained stable between 6 and 7% since 2001 (2). Among the MARPs in Uganda are fishing communities (FCs), which make up about 10% of the population and were until recently, due to their high mobility (3), believed to serve as reservoirs of infection for the general population (4,5). Several studies have indicated that HIV incidence in Ugandan FCs is up to 5 times, or more than 10 times depending on the community, higher than the national average, and therefore greatly contributes to the overall HIV burden in Uganda (6,7). This high incidence in FCs has been attributed to several factors including limited access to HIV prevention and treatment services as well as high-risk sexual behaviour (6), such as frequent partner change and low condom use (3,4).

Several HIV prevention interventions have been proven to effectively reduce HIV transmission. It has been shown that voluntary medical male circumcision (VMMC) decreases male susceptibility by 60%, while circumcised men living with HIV are as infectious as non-circumcised men (8). HIV counselling and testing (HCT) may lead to adoption of less risky behaviour, such as increased condom use and reduced sexual activity, as well as increased uptake of VMMC and oral pre-exposure prophylaxis (PrEP). On the other hand however, HIV-negative persons who repeatedly accept HCT are more likely to engage in high-risk sexual behaviour (9). Oral PrEP has been shown to reduce susceptibility by 67-75% (10), but high adherence to the prescribed doses is necessary for effective HIV prevention.

Another important intervention strategy is antiretroviral treatment (ART) for individuals living with HIV. In addition to prolonged life expectancy, persons living with HIV that are on ART have been shown to be less infectious (11). According to UNAIDS figures in 2017 (12), 68-77% of Ugandans living with HIV were receiving treatment. However, coverage in FCs is expected to be much lower than in the general population (13), because of barriers to treatment uptake such as stigma and inconvenience due to high mobility (14,15).

These HIV prevention strategies are however only partly effective and often not readily available to MARPs. It has been shown that minimal overlap of partly effective interventions increases the chances of synergistic effects (16). Cox et al. (17) found that using a combination of complementary interventions may be more practical for achieving substantial reductions in HIV incidence. Therefore, to achieve effective HIV control and have the greatest sustained impact on reducing new infections, several intervention strategies need to be combined to present a mix of effective behavioural, biomedical, and structural interventions. The Uganda national guidelines for HIV care incorporated the UNAIDS 90-90-90 (90% of people living with HIV aware of their status, 90% in care, and 90% virally suppressed) targets in 2016 (18). However, HIV prevention and treatment services may not be easily accessed by individuals living in FCs. Common barriers to accessing HIV prevention services include a lack of social support, stigma, long distances to healthcare facilities, and high population migration rates. Regarding HIV testing and ART, varied coverage was found among FCs but in general was not fulfilling the UNAIDS 95-95-95 targets in order to eliminate HIV by 2030. Hence to achieve global targets on HIV elimination, these FCs require special attention by implementing a combination of services tailored to the specific needs of this community (19).

Several modelling studies have shown the possible benefits of implementing combination prevention packages for reducing HIV incidence in high-risk populations (17,20,21). In the present study we investigated the impact of combining HIV prevention interventions on HIV dynamics in four FCs in Uganda. The interventions considered were VMMC, condom use, HCT, ART, and oral PrEP. The study findings provide additional knowledge on the use of targeted intervention combinations in the fight against the HIV epidemic.

## Methods

### Model description

We developed a population-level, deterministic, compartmental model of heterosexual HIV transmission incorporating different risk groups as is done in similar HIV transmission models (20,21). In our model the population aged 16-60 years is stratified into subgroups based on gender and sexual risk behaviour. The model consists of 14 compartments for females and 16 for males, representing susceptibility (with or without oral PrEP, and circumcision status for males) and treatment status (undiagnosed, diagnosed untreated, and diagnosed treated) in four disease stages. The rate at which susceptible individuals acquire HIV (i.e. force of infection) depends on their rate of partner change and PrEP/circumcision status, as well as their partner’s risk group, disease stage, and treatment status. It further depends on the type of partnership (by the frequency of coital acts) and probability of condom use per coital act (see Additional File 1). Once an individual is infected with HIV, they move through four disease stages: (i) an initial high viremia stage (acute infection), (ii) a low viremia stage (chronic infection), (iii) a pre-AIDS high viremia stage, and (iv) the final AIDS stage in which they remain until death from AIDS-related or other causes. It is assumed that individuals in the high viremia stages of infection are more infectious than those in the low viremia stage (22). Furthermore we assume that individuals in the untreated AIDS stage cease all sexual activity, while those in the treated AIDS stage greatly reduce their sexual activity.

Three sexual activity risk groups are defined: (i) low – individuals having only main partners, (ii) medium – individuals having main and non-main regular partners, and (iii) high – individuals having main, non-main regular, and casual partners. Hence an additional layer of heterogeneity is added by including different types of partnerships, with different associated risks, within each risk group. Although these partnerships are instantaneous in the model, the risk of infection throughout a partnership is a function of the frequency of coital acts per partnership, reflecting the duration and associated risk of a partnership (i.e. less coital acts indicates shorter duration but with a higher risk of infection due to a higher background HIV prevalence in these groups) (20). A detailed description of the model structure and parameters is presented in Figure S1 and Additional File 1. Below we describe the intervention measures.

### Target population

The target population of this study were four FCs around Lake Victoria, Uganda. Sociodemographic and prevalence data used to inform and calibrate the model came from the 2014 census and baseline serosurvey of the HIVCOMB study, respectively (23). Briefly, the HIVCOMB study was a pilot cluster-randomized trial assessing the feasibility of conducting combination HIV prevention interventions. Two rural and two urban FCs were included and in each urban-rural pair one cluster was assigned to receive the combination prevention package while the other cluster served as a control, receiving a standard prevention package. Before the community-wide intervention was introduced, a serosurvey was undertaken in a simple random sample from each community to determine HIV status. HIV prevalence estimates from this survey were used to calibrate our model to the actual situation in 2014.

### Model calibration

Most of the model parameters were unknown and had to be calibrated. Uncertainty ranges for all unknown parameters were constructed based on literature or, if unavailable, reasonably wide ranges were assumed (Table S2, see Additional File 1). Parameters that were calibrated included the transmission probability per coital act, parameters on baseline intervention uptake, and all behavioural parameters. The initial population size was set to 2000 and the proportion of individuals in each sexual risk group was fixed as those observed in the HIVCOMB survey: 33% of males and 43% of females in the low risk group, 7% of males and 2% of females in the medium risk group, 10% of males and 5% of females in the high risk group. The initial HIV prevalence was set to 1% in each risk group, with all of those in the initial high viremia stage.

The model was calibrated to the 2014 HIV prevalence in each sexual risk group, obtained from the 2014 HIVCOMB survey described above. Using Latin hypercube sampling (LHS) (24), 10 000 parameter input sets θ were sampled from the uncertainty ranges of the unknown parameters (Table S2, see Additional File 1). For each of these parameter sets, the model was run for 45 years (i.e. 1969-2014) to simulate an epidemic. Since we calibrated the model to only one time point, because no further data are available, overall goodness of fit (GOF) was measured as the sum of the squared deviations from the target prevalence in each risk group (25). An active learning approach, in which the parameter space is optimized in an iterative manner (26), was used to reduce uncertainty ranges for the unknown parameters by investigating the top 1% (based on GOF) of model outputs, Θ_*1%*_ (see Additional File 1).

### Interventions

HIV testing was assumed to have been available since 1987 (27). Persons living with HIV are diagnosed according to a rate of testing uptake that changes over time such that the proportion of infected individuals aware of their status increases linearly until reaching a specified level (21). When tested, infected individuals move to the ‘diagnosed’ compartment (representing awareness of their status) corresponding to their disease stage at that time. While diagnosed but untreated, individuals are assumed to slightly reduce their risk behaviour due to HIV awareness and/or counselling. Since results from studies about the effects of counselling on risk behaviour reduction are inconsistent (28), this was implemented as a conservative decrease of 10% in the number of sexual partners and a 10% increase in condom use.

ART was introduced in the pre-AIDS and AIDS stages from 2004 onward (29), and additionally in the other stages in 2011 (2). Depending on disease stage, a proportion of diagnosed individuals will initiate ART on average one year after diagnosis, and this proportion was assumed to increase over time. ART was assumed to reduce infectiousness by 96% (11). Treatment failure was included in the ART dropout rate which was set to an annual dropout of 20%. After ART dropout, individuals move back to the diagnosed untreated compartment and can re-initiate treatment at the same rate as treatment-naïve individuals. Individuals on ART were assumed to return to their baseline risk behaviour. ART and testing uptake were assumed to be lower in males compared to females (14).

Susceptible men opting for VMMC were assumed to have accepted HIV testing. Hence, VMMC uptake was modelled as a combination of the rate of testing uptake and the proportion of men getting circumcised on average one year after testing, which was assumed to increase over time. Circumcised males were assumed to have a relative susceptibility of 40% (8). Due to possible risk compensation, no reduction in sexual risk behaviour was assumed for circumcised males. We assumed that there was no VMMC before the introduction of HIV testing in 1987, although a constant proportion of males enter the population already circumcised (e.g. due to religion). The level of condom use was assumed to differ for each type of partnership, and to have increased over time until reaching a specified level.

Oral PrEP has been available since 2015 and hence was not included in the model calibration (2). Susceptible individuals initiating PrEP were assumed to have accepted HCT, and uptake was modelled as the combination of an annual probability of accepting HCT and a probability of initiating PrEP on average one year after testing, which was assumed to increase over time. Individuals on oral PrEP were assumed to have a relative susceptibility of 33% (10), further depending on the level of adherence which was assumed to be 60%. PrEP discontinuation was included as a fixed annual dropout of 20%. As for VMMC, no reduction in risk behaviour was assumed.

### Assessing the impact of combined interventions

As a baseline model, intervention uptake rates were kept constant for 15 years starting in 2014, although we slightly increased the proportion of individuals aware of their HIV-positive status to 65% to avoid zero rates of VMMC uptake. Since we had no information on the current level of oral PrEP uptake in these FCs but which is assumed to be very low, we did not include this in our baseline model.

The impact of intervention scale-up on HIV dynamics was investigated using the ‘best fit’ calibrated model. LHS was used to sample 500 input parameter sets θ_*I*_ from the uptake ranges of the intervention parameters (Table S3, see Additional File 1). To estimate long-term intervention impact, the calibrated model was run for an additional 15 years (i.e. 2014 – 2029). Starting in 2014, intervention uptake was gradually increased for 6 years (i.e. until 2020), after which a more steep increase was implemented for 9 years to mimic real-life combination prevention efforts. Different combinations of intervention uptake levels were then compared in terms of their impact on the relative reduction in HIV incidence and prevalence, AIDS-related mortality, and the total number of new infections, by comparing the model with scale-up of interventions to our baseline model without increased intervention uptake (i.e. ‘best fit’ intervention parameters as in Table S2, see Additional File 1).

### Uncertainty analysis

To account for the remaining uncertainty in the parameters of the ‘best fit’ calibrated model, which reflect only one possible epidemic trajectory, parameter ranges of simulated epidemics belonging to the top 1% of model outputs were also used to investigate the impact of combined interventions, resulting in 50 000 simulated scenarios (i.e. the 500 intervention parameter sets θ_*I*_ applied to 100 sets of model parameters Θ_*1%*_).

### Sensitivity analyses

We performed various sensitivity analyses to assess the influence of certain assumptions on the projected impact of intervention scale-up. In our best case scenario (i.e. calibrated model resulting in the highest reduction in incidence), we lowered the annual rate of PrEP discontinuation and ART dropout to 5%, increased the annual rate of ART dropout to 70%, assumed no increase in condom use after 2014, no PrEP use, PrEP adherence of 90%, and PrEP adherence of 30%. We also investigated the impact of equal HCT and ART uptake in males and females.

## Results

### Model calibration

Tables 1 and 2 show the HIV prevalence estimates and intervention coverage levels, respectively, from the ‘best fit’ model after completing calibration. The corresponding model parameters are shown in Table S2 (see Additional File 1). HIV dynamics in terms of prevalence and incidence in the different sexual risk groups for the baseline model (i.e. without intervention scale-up) are shown in Figure S3 (see Additional File 1). It can be seen that even without increasing the intervention uptake, HIV prevalence and incidence would already decline after 2014 (overall, a decline of 19.89% in prevalence and 30.50% in incidence when comparing 2029 to 2014), with steeper declines observed for females in the medium and high risk groups.

**Table 1.** Prevalence (%) observed in the HIVCOMB study and estimates of prevalence (%) and incidence (per 100 PY) from the ‘best fit’ model (based on GOF) after completing model calibration.

|  | **Observed prevalence (95% CI)** |  | **Prevalence (95% CI) best fit model** |  | **Incidence (per 100 PY) best fit model** |  |
| --- | --- | --- | --- | --- | --- | --- |
| **Total** | 18.13 (15.35 – 20.91) |  | 17.65 (16.48 – 18.85) |  | 2.69 |  |
| **Risk group** | **Male** | **Female** | **Male** | **Female** | **Male** | **Female** |
| Low risk | 11.74 (8.30 – 16.35) | 20.89 (16.77 – 25.70) | 13.76 (12.05 – 15.60) | 19.72 (17.86 – 21.65) | 1.67 | 2.55 |
| Medium risk | 20.83 (11.73 – 34.26) | 33.33 (15.18 – 58.29) | 20.87 (15.91 – 26.44) | 33.51 (21.52 – 48.27) | 2.50 | 4.57 |
| High risk | 14.29 (8.17 – 23.80) | 36.11 (22.48 – 52.43) | 13.84 (10.67 – 17.74) | 36.64 (27.97 – 46.62) | 1.50 | 5.02 |

**Table 2.** Intervention coverage (%) in 2014 for the calibrated ‘best fit’ model.

|  | **Aware of HIV+ status** | **On ART of all HIV infected** | **On ART of those aware of HIV+ status** | **Circumcised** |
| --- | --- | --- | --- | --- |
| **Overall** | 60.92% | 14.81% | 24.31% | - |
| **Female** | 65.36% | 17.29% | 26.45% | - |
| **Male** | 55.19% | 11.62% | 21.05% | 15.29% |

### Impact of combined interventions

Using the parameters from the ‘best fit’ calibrated model, the intervention uptake levels shown in Table S3 (highest impact scenario; see Additional File 1) resulted in a relative reduction in HIV incidence of 86.68% at the end of 15 years of intervention scale-up, compared to a baseline scenario without scale-up. The corresponding coverage after 15 years for each of the interventions is shown in Table 3 (see Figure S4 in Additional File 1 for coverage levels over time), and the relative reduction in prevalence, incidence, AIDS-related mortality, and number of new infections per risk group are shown in Table 4. Overall incidence after 15 years was 1.87 per 100 PY in the baseline scenario compared to 0.25 per 100 PY after intervention scale-up. The relative reduction in incidence was highest for males in the low risk group (88.15%) and lowest for females in the high risk group (83.15%). In each risk group, relative reductions were slightly greater for males.

**Table 3.** Intervention coverage that had the highest impact on incidence after 15 years of scale-up. Coverage over time is shown in Figure S4 e Additional File 1).

|  | Aware of HIV+ status | On ART of all HIV infected | On ART of those aware of HIV+ status | Circumcised | Oral PrEP |
| --- | --- | --- | --- | --- | --- |
| Overall | 87.13% | 65.28% | 74.93% | - | 2.65% |
| Female | 89.40% | 68.92% | 77.09% | - | 3.16% |
| Male | 83.87% | 60.09% | 71.65% | 22.58% | 2.24% |

**Table 4.** Highest intervention impact estimates of HIV prevalence (%) without (baseline) and with scale-up of interventions; relative reduction in prevalence, incidence, AIDS-related mortality, and number of new infections over 15 years (obtained by comparing the model with intervention scale-up to the baseline model after 15 years). LR = low risk, MR = medium risk, HR = high risk.

|  | Baseline | Intervention scale-up | Relative reduction (compared to baseline) |  |  |  |
| --- | --- | --- | --- | --- | --- | --- |
|  |  |  | Prevalence | Incidence | AIDS-related mortality | New infections over 15 years |
| **Overall** | **14.14% (13.17 – 15.13)** | **8.75% (7.98 – 9.55)** | **0.3810** | **0.8668** | **0.5376** | **0.4123** |
| **Female LR** | 16.78% (15.18 – 18.42) | 10.51% (9.26 – 11.89) | 0.3739 | 0.8672 | 0.5478 | 0.4092 |
| **Female MR** | 27.90% (16.14 – 40.96) | 18.34% (9.87 – 31.41) | 0.3425 | 0.8500 | 0.5320 | 0.3848 |
| **Female HR** | 29.85% (21.82 – 39.42) | 20.33% (13.26 – 28.28) | 0.3192 | 0.8315 | 0.5170 | 0.3651 |
| **Male LR** | 10.88% (9.50 – 12.35) | 6.37% (5.33 – 7.55) | 0.4144 | 0.8815 | 0.5389 | 0.4299 |
| **Male MR** | 15.67% (11.81 – 20.52) | 9.64% (6.59 – 13.63) | 0.3848 | 0.8651 | 0.5222 | 0.4104 |
| **Male HR** | 9.53% (7.12 – 12.45) | 6.07% (4.15 – 8.48) | 0.3636 | 0.8443 | 0.5056 | 0.3983 |

Figure 1 shows the evolution of HIV incidence and prevalence in the total population for the baseline model and the model with intervention scale-up. After intervention scale-up, a much steeper decline in prevalence and incidence is observed. The results also indicate that implementation of combined interventions would yield a total of 408 infections averted over 15 years (989 new infections under the baseline scenario, vs. 581 new infections under the highest impact scale-up scenario).

**Figure 1.**
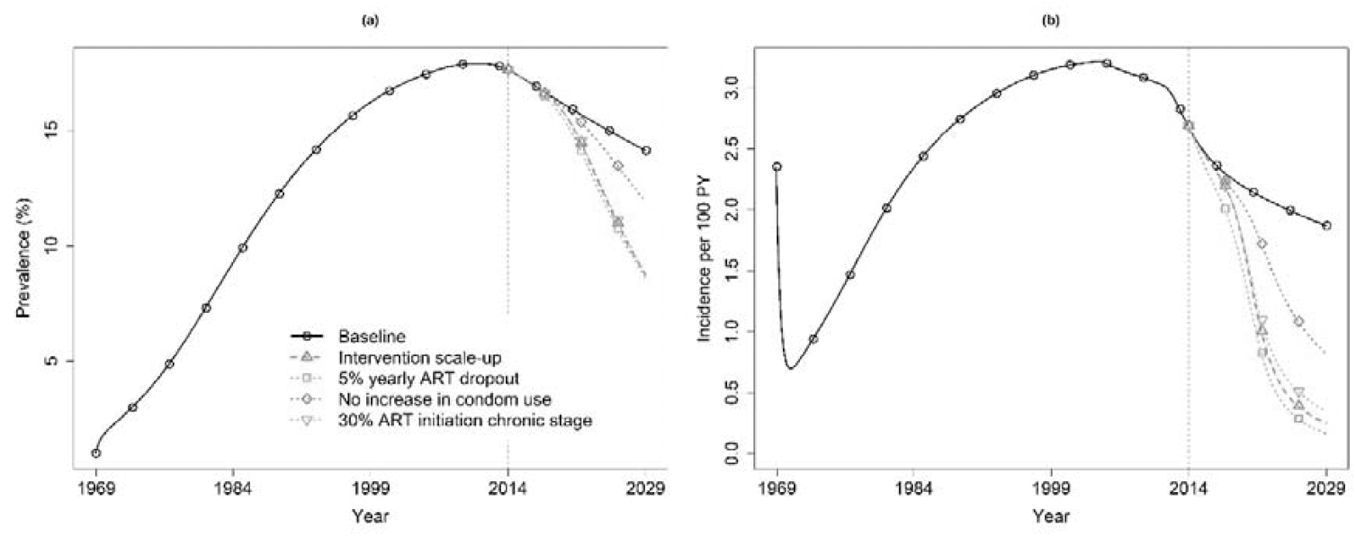
Overall HIV prevalence (a) and incidence (b) obtained under the baseline model (full black line) and the ‘best fit’ model with intervention scale-up (dashed red line) giving the highest reduction in incidence, 1969-2029. Intervention scale-up started in 2014 (dotted grey vertical line). The impact of decreasing the yearly rate of ART dropout, assuming no increase in condom use, and lowering the probability of ART initiation in the chronic stage are also shown. Parameter estimates for the baseline model are shown in Table S2, while parameter estimates for the intervention uptake under the highest-impact scenario are shown in Table S3 (see Additional File 1).

### Uncertainty analysis

When accounting for parameter uncertainty, the relative reductions in incidence ranged from 30.90% to 86.80% (median 65.68%, IQR 55.48-76.66%). The relative reductions in prevalence ranged from 10.05% to 38.66% (median 26.24%, IQR 20.17-32.02%). Figure 2 shows that condom use in the low-risk group, ART initiation during the chronic stage, and the proportion of diagnosed individuals confer the greatest impact on the predicted relative reduction in HIV incidence, with higher uptake levels leading to higher reductions (Figure 3), when implemented in combination with the other interventions.

**Figure 2.**
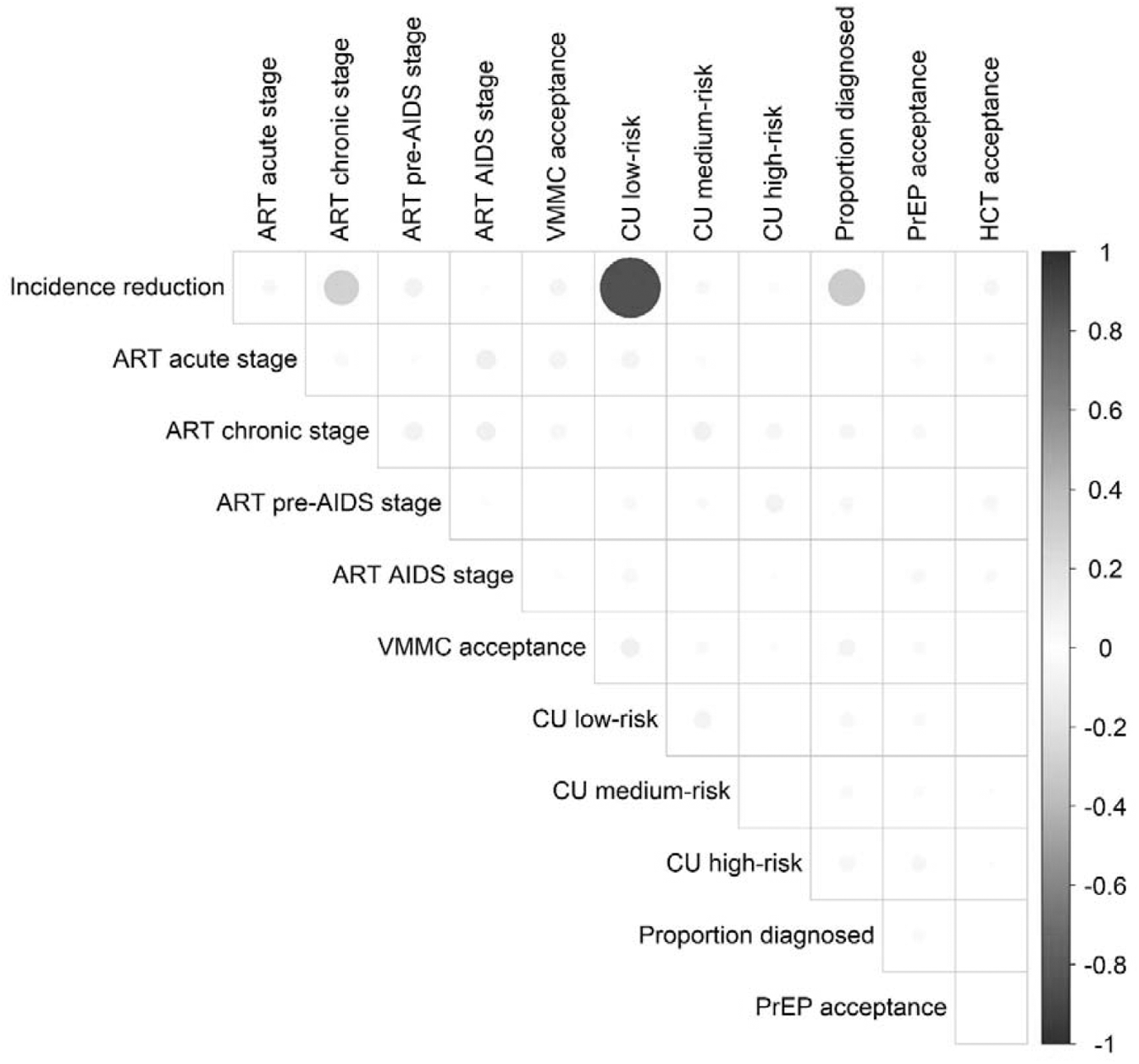
Correlation between each intervention component and the relative reduction in HIV incidence based on the 50 000 combinations used in the uncertainty analysis.

**Figure 3.**
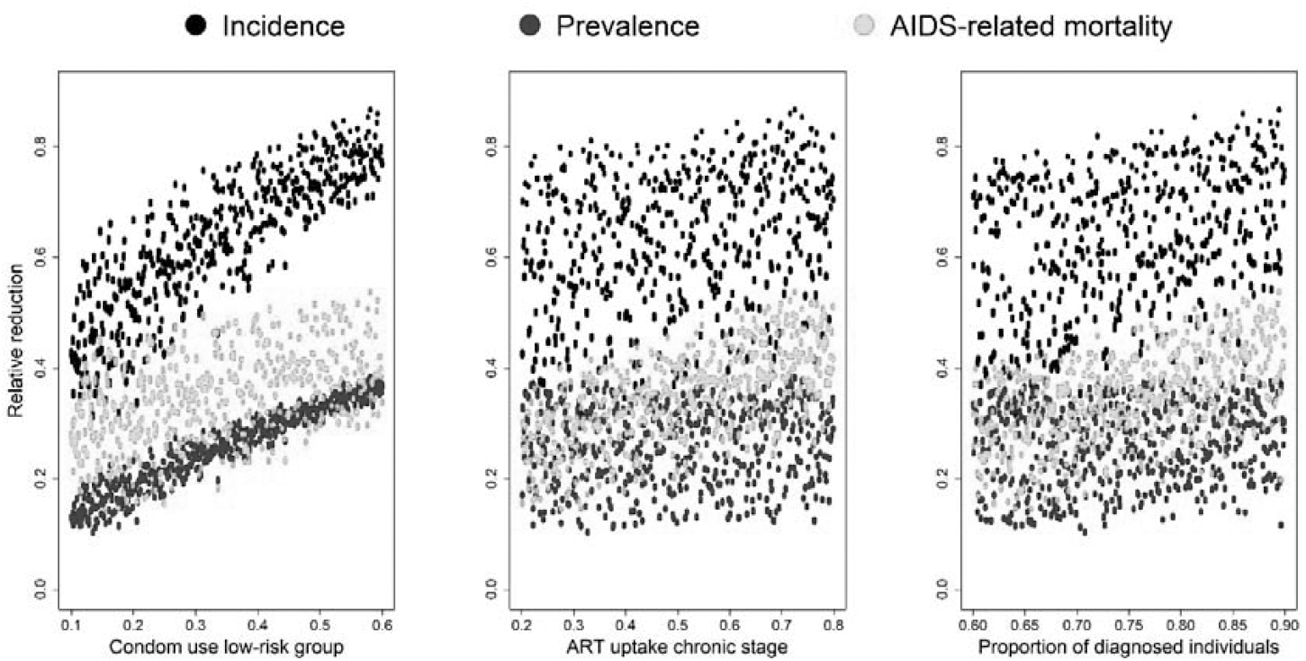
Relative reduction in incidence, prevalence, and AIDS-related mortality for the three most important intervention components (condom use in the low risk group, ART initiation during the chronic stage, and the proportion of diagnosed individuals).

### Sensitivity analysis

Results of the sensitivity analyses are shown in Table S4 (see Additional File 1) and Figure 1 (selected scenarios). Scenarios varying the assumptions regarding oral PrEP use did not affect the projected impact of the interventions, probably due to the low coverage levels. Assuming equal testing and treatment uptake in males and females slightly increased the projected reduction in incidence from 86.68% to 88.11%. Reducing the annual rate of ART dropout from 20% to 5% resulted in an increase of the projected reduction in incidence from 86.68% to 91.16%. Under this scenario, a substantially greater reduction in AIDS-related mortality was seen, as well as in the number of new infections over 15 years. In contrast, increasing the annual rate of ART dropout to 70% resulted in a decrease of the projected reduction in incidence from 86.68% to 77.78%, as well as a substantial decrease of the projected reduction in AIDS-related mortality and number of new infections over 15 years. Assuming no increase in condom use after 2014 had a large impact, with the relative reduction in incidence dropping to 56.19%. To assess the impact of early ART initiation, we lowered the probability of initiating ART in the chronic stage to 30%. This resulted in 81.54% reduction in incidence and 41.54% reduction in AIDS-related mortality (Table S4, see Additional File 1). In this scenario, only 55.94% of those aware of their HIV status were on ART, compared to 74.93% on ART in the highest impact scenario that assumed a 72.51% probability of initiating ART in the chronic stage.

## Discussion

Results of our modelling study have indicated that we can achieve as high as 87% relative reduction in HIV incidence through combination prevention. We have projected that this reduction in incidence can be achieved by 2029 using the following combination of intervention coverage levels: (i) condom use in about 60% of sexual acts, (ii) 23% of susceptible men being circumcised, (iii) 87% of people living with HIV aware of their status, 75% of those aware of their status on ART, and (v) about 3% of susceptible individuals on PrEP. In our best case scenario, reaching the above coverage levels, we projected that HIV incidence may reduce to 2.5 per 1000 PY, and overall prevalence to 8.75%. These numbers are still higher than for the general population in Uganda, and incidence is still above the threshold of 1 per 1000 PY that can be seen as HIV elimination (30). It should be noted that other combinations of coverage levels of the intervention components may lead to similar reductions in incidence. The decline in prevalence and incidence observed in our baseline model is in line with the globally declining number of new HIV infections since 1996 (1).

Our projected coverage levels are modest in comparison to the Uganda national targets and hence attainable. For instance the national target for ART coverage by 2020 was 80% (31). Moreover, the Uganda AIDS Commission laid down strategies to improve HIV testing and linkage to care in FCs by partnering with the Ministry of Agriculture and Fisheries (31). The Uganda government in her HIV and AIDS Strategic Plan 2015/2016-2019/2020 emphasized the need to scale up combination prevention strategies to critical levels by 2020 (2). Our projected levels are well within these 2020 national targets. The prevention components we considered in our study are key in objective two of this National Strategic Plan.

Our projections indicate that condom use and ART will have the largest impact on HIV transmission in these FCs. Without an increase in condom use, the projected reduction in incidence was only 56.19% compared to our baseline scenario. Sensitivity analysis also showed that initiating ART early plays an important role in achieving substantial reductions in incidence and AIDS-related mortality. Lowering the probability of ART initiation during the chronic stage decreased the projected impact to an incidence reduction of 81.54%. A previous modelling study in a hyperendemic setting with relatively low levels of condom use found that increasing early ART coverage rather than providing PrEP to uninfected individuals would be more cost-effective, but that early ART alone is not sufficient for reducing HIV incidence to very low levels (20). In our projections, early ART did have a substantial impact on reductions in HIV incidence, although combined with relatively high levels of condom use. Uncertainty analyses using 50 000 possible scenarios also showed that condom use in the low risk group was the most important component of the prevention package. As it is known that achieving high levels of condom use in these FCs is difficult, public health initiatives should focus on adequate ART delivery in this hard-to-reach population and ensure sufficient adherence, combined with increased efforts regarding condom promotion and distribution.

A systematic review in 2020 found that fisher folk have a preference for PrEP and VMMC over other prevention services such as condom use, and that there is an urgent need for innovative approaches to HIV care delivery in FCs (19). In our projections, oral PrEP use did not seem to play a very important role in reducing HIV incidence, but coverage in the best case scenario was very low (around 3%). However, increasing oral PrEP uptake is expected to further reduce HIV incidence. Oral PrEP uptake and adherence to a daily regimen have not been optimal in high-risk populations (32). This may be mitigated by the use of long-acting injectables that do not require a daily regimen (33), although a previous study has found that fisher folk would prefer oral PrEP (34). In addition, several barriers to PrEP initiation such as travel distance to health facilities and community stigma should be overcome first (35). Furthermore, efforts should be made to avoid risk-compensating behaviour in susceptible individuals on PrEP, which could cancel out its effect.

An advantage of the compartmental model used in this study is that it accounts for a large part of the heterogeneity in HIV risk behaviour through the inclusion of different sexual risk groups that can form different types of partnerships, each with its own associated risk. An alternative approach would be to use an individual-based model (IBM) which can incorporate a higher degree of heterogeneity at the individual level, such as more detailed information on sexual mixing behaviour (35). When model structure becomes too complex to capture in a compartmental modelling framework, an IBM may be the solution. In addition, when using an IBM, intervention policies can be tailored to the individual, but this was beyond the scope of the present study and would require more detailed data.

This study has several limitations. First, the model was calibrated to observed HIV prevalence at only one time point, since these were the only data available. Although the ‘best fit’ model represents only one possible epidemic trajectory, we performed an uncertainty analysis using a range of model parameters. Second, the model does not explicitly account for other STIs that are known to be co-factors in HIV infection. These data were not available from the HIVCOMB study, and it was not the focus in this work to investigate the contribution of other STIs. However, since the model was calibrated to the observed prevalence data, we have assumed that it does implicitly account for the role of other STIs in HIV transmission. Third, the model does not account for behavioural change; we assumed that people stay in the same sexual risk group over time. Fourth, the model does not incorporate stochasticity, which could provide model flexibility to accommodate changes in the transmission rates that might occur due to unobserved stochastic processes (36). We also did not account for the fact that some interventions may induce risk compensating behaviour such as reduced condom use and more sexual risk-taking. In addition, we have not addressed the complex relationship between alcohol use, intimate partner violence (IPV), and HIV infection. IPV may increase a woman’s risk of HIV infection, for example due to forced unprotected sexual intercourse or because of their lack of power to negotiate condom use (37). Lastly, our model was calibrated to prevalence data due to the lack of incidence data. However, although prevalence is influenced by new infections as well as deaths, we also assessed the impact of the combined interventions on incidence.

## Conclusions

Despite the highlighted limitations, we have projected that a substantial reduction in HIV incidence can be achieved through combination prevention without necessarily reaching high coverage of all the specific intervention package components. Although Ugandan FCs may have extensive sexual networks bridging into the general population, Ratmann et al. (5) have recently reported that only 1.3% of HIV transmissions occurred from lakeside to inland areas. However, HIV transmission from inland to lakeside areas was slightly higher (3.7%), hence future work could incorporate partnerships formed outside of the FCs, to investigate the interplay between reducing HIV prevalence in either inland or lakeside areas (4). It should be noted that the projections obtained in the present study are specific to the included Ugandan FCs and may not necessarily be generalizable to other settings with a different underlying HIV incidence or epidemic stage, as for instance the general population in Uganda.

## Data Availability

R code accompanying this manuscript is available on GitHub.

https://github.com/cecilekremer/hivug

## List of abbreviations

AIDS: Acquired Immunodeficiency Syndrome
ART: Antiretroviral therapy
FC: Fishing community
GOF: Goodness of fit
HCT: HIV counselling and testing
HIV: Human Immunodeficiency Virus
IPV: Intimate partner violence
LHS: Latin hypercube sampling
MARP: Most at risk population
PrEP: Pre-exposure prophylaxis
STI: Sexually transmitted infection
VMMC: Voluntary medical male circumcision

## Declarations

### Ethics approval and consent to participate

The HIVCOMB study was approved by the Uganda Virus Research Institute, the Uganda National Council of Science and Technology, and the London School of Hygiene and Tropical Medicine ethics committees.

### Consent for publication

Not applicable.

### Availability of data and materials

All data generated or analysed during this study are included in this published article and its supplementary information files. Computer code accompanying this manuscript is available at https://github.com/cecilekremer/hivug.

### Authors’ contributions

RNN, NH, and CK contributed to conceptualization of the study. RNN, AK, MK, and JS were involved in the HIVCOMB study. CK and RNN contributed to the analysis code. CK performed the analysis. NH, RNN, CK, AK, MK, and JS contributed to the interpretation of results. CK and RNN drafted the manuscript. All co-authors critically reviewed and revised the manuscript.

## Acknowledgements

Project carried out with the support of the Fund AIDS managed by the King Baudouin Foundation. The computational resources and services used in this work were provided by the VSC (Flemish Supercomputer Center), funded by the Research Foundation Flanders (FWO) and the Flemish Government department EWI. The authors would like to thank Helen A. Weiss (LSHTM) for her insightful comments on an earlier draft of this manuscript.

## Additional material

**Additional File 1. Detailed model description**.

**Figure S1. Model structure for males in risk group *j* (which is the same for females, without circumcision compartments)**.

**Figure S2. Parameters that are highly informed by the data**. Dashed lines represent the density *f*_U_ of the uniform distribution on the uncertainty range of the input parameter. Solid lines represent the density *f*_Θ1%_ corresponding to the values within the subspace Θ_*1%*_. A peaked unimodal density *f*_Θ1%_ indicates that the parameter was highly informed by the data.

**Figure S3. HIV transmission dynamics**. HIV prevalence (a) and incidence (b) in the different sexual risk groups obtained from the ‘best fit’ model. Filled symbols on the dotted grey vertical line represent the observed prevalence in 2014 obtained from the HIVCOMB study, to which the model was calibrated.

**Figure S4. Intervention coverage over time**. (a) HIV infected individuals on ART, (b) HIV infected individuals aware of their status on ART, (c) circumcised susceptible males, and (d) individuals aware of their HIV+ status. Dashed lines indicate the year 2011 when ART was implemented in the chronic stage, dotted lines indicate the year 2020 when intervention scale-up was started, dash-dotted lines indicate 1987 when HIV testing was introduced.

**Table S1. Probability of transmission per coital act**. Baseline corresponds to the chronic low viral load stage.

**Table S2. Uncertainty ranges and fixed values for the baseline model parameters and value in the ‘best fit’ solution**.

**Table S3. Uptake ranges for the intervention parameters and their values in the highest impact scenario**.

**Table S4. Sensitivity analyses for the impact of combined interventions**.

