## Supplementary Material for "Modelling the impact of combining HIV prevention interventions on HIV dynamics in fishing communities in Uganda"

### Additional File 1: Detailed description of the compartmental model - Supplementary Tables and Figures

#### Model description

We developed a population-level, deterministic, compartmental model of heterosexual HIV transmission in the population aged 16-60 years, stratified by gender ($r$) and sexual risk group ($j$). Susceptible individuals are either not receiving PrEP ($S^1$) or receiving PrEP ($S^2$), with additional compartments for circumcised males (not receiving PrEP, $S^3$, or receiving PrEP, $S^4$). A proportion $p_c$ of males is assumed to enter the susceptible population already circumcised. Once infected, individuals are assumed to move through four stages of disease progression: (i) an initial high viral (HV) load stage ($H$), (ii) a low viral (LV) load stage ($Y$), (iii) a pre-AIDS high VL stage ($Z$), and (iv) a final AIDS stage ($A$). When tested (according to a rate of diagnosis $\tau^T$ such that the proportion of infected individuals aware of their status increases linearly over time), infected individuals move to the corresponding 'diagnosed' compartment (representing awareness of HIV+ status) in which they are assumed to (slightly) reduce their risk behaviour due to HIV counseling. Infected individuals that have been tested and hence are aware of their disease status initiate ART with a certain probability $p_{ART}$ (increasing over time) depending on their disease stage. ART dropout is also modelled and includes treatment failure (i.e., individuals move back to the untreated compartment corresponding to their disease stage). Individuals that have dropped out of ART may then restart treatment with the same probability as previously untreated individuals. Susceptible individuals that have been tested but are not infected initiate PrEP with a certain probability $p_{PrEP}$ that is increasing over time from 2015 onwards [1]. PrEP discontinuation $\omega$ and ART dropout $\gamma$ are modeled as fixed annual rates. Figure S1 displays the model structure for males in risk group $j$. For females, the model structure is equivalent but without the leftmost 'circumcised' compartments. We assume HIV testing to be available since 1987 [2], and

ART since 2004 for individuals in the (pre-)AIDS stage [3] and since 2011 for all stages [1]. Intervention uptake in males is assumed to be lower than in females [4].
The model parameters and their uncertainty ranges are shown in Table S2. Let subscripts $r'$ and $p$ denote the gender and risk group of a partner of an individual from subgroup $(r, j)$. The model is described by the following set of ordinary differential equations (for females), where $N_{rj}$ is the total number of individuals in subgroup $(r, j)$ and $\pi^x_{rj}$ is the force of infection.

$$\frac{dS^1_{rj}}{dt} = (\mu + \psi)N_{rj} + \omega S^2_{rj} - (\pi^1_{rj} + \theta + \psi)S^1_{rj}$$

$$\frac{dS^2_{rj}}{dt} = \theta S^1_{rj} - (\pi^2_{rj} + \omega + \psi)S^2_{rj}$$

$$\frac{dH^1_{rj}}{dt} = \pi^1_{rj}S^1_{rj} + \pi^2_{rj}S^2_{rj} - (\nu^1 + \tau^T + \psi)H^1_{rj}$$

$$\frac{dH^2_{rj}}{dt} = \tau^T H^1_{rj} + \gamma H^3_{rj} - (\nu^1 + \tau^A_1 + \psi)H^2_{rj}$$

$$\frac{dH^3_{rj}}{dt} = \tau^A_1 H^2_{rj} - (\nu^2 + \gamma + \psi)H^3_{rj}$$

$$\frac{dY^1_{rj}}{dt} = \nu^1 H^1_{rj} - (\Omega^1 + \tau^T + \psi)Y^1_{rj}$$

$$\frac{dY^2_{rj}}{dt} = \nu^1 H^2_{rj} + \tau^T Y^1_{rj} + \gamma Y^3_{rj} - (\Omega^1 + \tau^A_2 + \psi)Y^2_{rj}$$

$$\frac{dY^3_{rj}}{dt} = \nu^2 H^3_{rj} + \tau^A_2 Y^2_{rj} - (\Omega^2 + \gamma + \psi)Y^3_{rj}$$

$$\frac{dZ^1_{rj}}{dt} = \Omega^1 Y^1_{rj} - (\Psi^1 + \tau^T + \psi)Z^1_{rj}$$

$$\frac{dZ^2_{rj}}{dt} = \Omega^1 Y^2_{rj} + \tau^T Z^1_{rj} + \gamma Z^3_{rj} - (\Psi^1 + \tau^A_3 + \psi)Z^2_{rj}$$

$$\frac{dZ^3_{rj}}{dt} = \Omega^2 Y^3_{rj} + \tau^A_3 Z^2_{rj} - (\Psi^2 + \gamma + \psi)Z^3_{rj}$$

$$\frac{dA^1_{rj}}{dt} = \Psi^1 Z^1_{rj} - (\delta^1 + \tau^T + \psi)A^1_{rj}$$

$$\frac{dA^2_{rj}}{dt} = \Psi^1 Z^2_{rj} + \tau^T A^1_{rj} + \gamma A^3_{rj} - (\delta^1 + \tau^A_4 + \psi)A^2_{rj}$$

$$\frac{dA^3_{rj}}{dt} = \Psi^2 Z^3_{rj} + \tau^A_4 A^2_{rj} - (\delta^2 + \gamma + \psi)A^3_{rj}$$

For males, the equations below differ from those for females due to the inclusion of circumcision compartments.

$$\frac{dS^1_{rj}}{dt} = (1 - p_c)(\mu + \psi)N_{rj} + \omega S^2_{rj} - (\pi^1_{rj} + \sigma + \theta + \psi)S^1_{rj}$$

$$\frac{dS^2_{rj}}{dt} = \theta S^1_{rj} - (\pi^2_{rj} + \sigma + \omega + \psi) S^2_{rj}$$

$$\frac{dS^3_{rj}}{dt} = p_c(\mu + \psi) N_{rj} + \sigma S^1_{rj} + \omega S^4_{rj} - (\pi^3_{rj} + \theta + \psi) S^3_{rj}$$

$$\frac{dS^4_{rj}}{dt} = \theta S^3_{rj} + \sigma S^2_{rj} - (\pi^4_{rj} + \omega + \psi) S^4_{rj}$$

$$\frac{dH^1_{rj}}{dt} = \pi^1_{rj} S^1_{rj} + \pi^2_{rj} S^2_{rj} + \pi^3_{rj} S^3_{rj} + \pi^4_{rj} S^4_{rj} - (\nu^1 + \tau^T + \psi) H^1_{rj}$$

#### Force of infection

Individuals can form three different types of partnerships: (i) main partnerships (can be formed within or between all risk groups), (ii) non-main regular partnerships (can be formed within or between medium and high risk groups), and (iii) casual partnerships (can be formed only within the high risk group). In all of the following notation, $j$ denotes a person's risk group, $p$ denotes the risk group of the partner, and $i$ denotes the type of partnership. The **force of infection,** $\pi^x_{rj}$ (i.e. the probability that a susceptible acquires infection from their sexual partnerships, per unit time), is frequency-dependent and is defined separately for each subgroup, and for susceptibles not on PrEP ($x = 1$ for females/uncircumcised males, and $x = 3$ for circumcised males) and susceptibles on PrEP ($x = 2$ for females/uncircumcised males and $x = 4$ for circumcised males).

$$\pi^x_{rj} = \sum_{r'} \sum_{p} \sum_{i} \left[ d_{rji} \rho_{rjpi} \sum_{s} \sum_{h} \left( \frac{I^{s,h}_{r'p}}{N_{r'p}} \varphi^x_{rjr'pish} \right) \right]$$

where

$\varphi^x_{rjr'pish}$ is the probability of transmission for a susceptible in subgroup $(r, j)$ having a sexual partnership of type $i$ with an individual from subgroup $(r', p)$ who is in disease stage $s$ (i.e. probability of infection per partnership per unit time), and depends on PrEP use/circumcision status ($x$) of the susceptible as well as ART/HCT status ($h$) of the infected individual

$d_{rji}$ is the rate at which individuals in subgroup $(r, j)$ form partnerships of type $i$ with all other groups, per unit time

$\rho_{rjpi}$ is the probability that individuals in subgroup $(r, j)$ form sexual partnerships of type $i$ with individuals in subgroup $(r', p)$ $\rightarrow$ mixing parameter (controls the rate of partnership formation between specific subgroups)

$\frac{I^{s,h}_{r'p}}{N_{r'p}}$ is the proportion of individuals in risk group $(r', p)$ that are infected and in disease stage $s$ with ART/HCT status $h$

Then, $d_{rji}\rho_{rjpi}$ represents the total number of partnerships of type $i$ that an individual from $(r, j)$ forms with those from $(r', p)$, per unit time. A correction for the resulting imbalance in partnerships is described below.

The **mixing parameters** $\rho_{rjpi}$ represent the extent to which individuals within any behavioral subgroup form sexual partnerships of type $i$ with individuals from different subgroups. Sexual mixing is assumed to be proportionate but with a tendency for within-class mixing,

$$\rho_{rjpi} = (1 - \mathcal{A}_i)\frac{d_{r'pi}n_{r'p}}{\sum_{\forall l} d_{r'li}n_{r'l}} + \mathcal{A}_i\zeta_{jp}$$

where

$\mathcal{A}_i$ is the degree of assortativity for sexual partnerships of type $i$

- $\mathcal{A}_i$ is assumed to be closer to 1 (i.e. assortative mixing) for main partnerships, and closer to 0 (i.e. random mixing) for non-main regular partnerships. For casual partnerships, mixing is always assortative ($\mathcal{A}_{casual} = 1$) since these can only be formed with individuals from subgroup $(r', 3)$.

$\zeta_{jp}$ is 1 if $j = p$ and 0 otherwise

$n_{rj}$ is the total number of individuals in each subgroup

Hence, for main and non-main regular partnerships, a proportion $\mathcal{A}_i$ of partnerships are formed only with individuals from the same risk group. All others are formed with each risk group, proportional to the number of such partnerships offered by those risk groups.

The $\rho_{rjpi}$ are defined for males and females from each sexual risk group, and for each type of partnership, e.g. $\rho_{F32_{reg}}$ denotes the level of mixing for non-main regular partnerships between females from the high risk group with males from the medium risk group. In order to balance the number of partnerships between behavioral subgroups, the rates of partner acquisition ($d_{rji}$) are adjusted such that they will depend also on the behavioral subgroup of the partner (denoted by $p$) [5]. The degree of initial imbalance can be obtained as

$$B_{jpi} = \frac{d_{r'pi}\rho_{r'pji}n_{r'p}}{d_{rji}\rho_{rjpi}n_{rj}}$$

and equals 1 if the number of partnerships is balanced. $B_{jpi}$ then represents the discrepancy in number of partnerships (of type $i$), which can be corrected by altering the partner acquisition rates of one or both sexes such that

$$d_{r'pji} = d_{r'pi}B_{jpi}^{x_i - 1}$$

and

$$d_{rjpi} = d_{rji} B_{jpi}^{x_i}$$

where $x_i$ is a constant controlling the extent to which each gender compromises with the wishes of the other gender (calibrated for each type of partnership). For example if both sexes were to compromise equally, $x_i = 0.5$. In this way, the following constraint on sexual mixing is met

$$d_{rjpi} \rho_{rjpi} N_{rj} = d_{r'pji} \rho_{rpji} N_{r'p}$$

The **transmission parameter** $\varphi^x_{rjr'pish}$ denotes the probability of transmission per partnership per unit time and depends on the susceptible individual's risk group ($j$) and PrEP/circumcision status ($x$), and the partner's risk group ($p$), disease stage ($s$) and HCT/ART use ($h$). It further depends on the type of partnership by the frequency of coital acts ($\eta_{jpi}$), probability of condom use per coital act ($f_i$), and efficacy of condom use ($e_c$). Condom use was modeled among the entire population, regardless of HIV status. The probability of transmission per coital act $\beta_r^{s,h,x}$ is defined in terms of the efficacy of ART ($e_A$), PrEP efficacy ($e_P$), level of PrEP adherence ($P_r$), efficacy of circumcision ($e_{circ}$), and the infected partner's disease stage (by a multiplication factor $\alpha$). Hence, assuming a baseline $\beta_r^{1,1,1}$ for individuals in the chronic stage of infection that are not on ART/HCT and when the susceptible is not on PrEP, we obtain the following probabilities of transmission (see Table S1).

$$\beta_r^{2,1,1} = \alpha_1 \beta_r^{1,1,1}$$
$$\beta_r^{3,1,1} = \alpha_2 \beta_r^{1,1,1}$$
$$\beta_r^{4,1,1} = 0$$
$$\beta_r^{1,3,1} = \beta_r^{1,1,1}(1 - e_A)$$
$$\beta_r^{2,3,1} = \beta_r^{1,1,1}(1 - e_A)$$
$$\beta_r^{3,3,1} = \beta_r^{1,1,1}(1 - e_A)$$
$$\beta_r^{4,3,1} = \beta_r^{1,1,1}(1 - e_A)$$

$$\beta_r^{1,1,2} = \beta_r^{1,1,1}(1 - e_P P_r)$$
$$\beta_r^{2,1,2} = \beta_r^{2,1,1}(1 - e_P P_r)$$
$$\beta_r^{3,1,2} = \beta_r^{3,1,1}(1 - e_P P_r)$$
$$\beta_r^{4,1,2} = 0$$
$$\beta_M^{1,1,3} = \beta_M^{1,1,1}(1 - e_{circ})$$
$$\beta_M^{2,1,3} = \beta_M^{2,1,1}(1 - e_{circ})$$
$$\beta_M^{3,1,3} = \beta_M^{3,1,1}(1 - e_{circ})$$
$$\beta_M^{4,1,3} = 0$$

$$\beta_r^{1,3,2} = \beta_r^{1,3,1}(1 - e_P P_r)$$
$$\beta_r^{2,3,2} = \beta_r^{2,3,1}(1 - e_P P_r)$$
$$\beta_r^{3,3,2} = \beta_r^{3,3,1}(1 - e_P P_r)$$
$$\beta_r^{4,3,2} = \beta_r^{4,3,1}(1 - e_P P_r)$$
$$\beta_M^{1,3,3} = \beta_M^{1,3,1}(1 - e_{circ})$$
$$\beta_M^{2,3,3} = \beta_M^{2,3,1}(1 - e_{circ})$$
$$\beta_M^{3,3,3} = \beta_M^{3,3,1}(1 - e_{circ})$$
$$\beta_M^{4,3,3} = \beta_M^{4,3,1}(1 - e_{circ})$$

$$\beta_M^{1,3,4} = \beta_M^{1,3,3}(1 - e_P P_M)$$
$$\beta_M^{2,3,4} = \beta_M^{2,3,3}(1 - e_P P_M)$$
$$\beta_M^{3,3,4} = \beta_M^{3,3,3}(1 - e_P P_M)$$
$$\beta_M^{4,3,4} = \beta_M^{4,3,3}(1 - e_P P_M)$$
$$\beta_M^{1,1,4} = \beta_M^{1,1,3}(1 - e_P P_M)$$
$$\beta_M^{2,1,4} = \beta_M^{2,1,3}(1 - e_P P_M)$$
$$\beta_M^{3,1,4} = \beta_M^{3,1,3}(1 - e_P P_M)$$
$$\beta_M^{4,1,4} = 0$$

Then, the transmission parameters $\varphi^x_{rjr'pish}$ are as follows

- To women/uncircumcised men not on PrEP:

$$\varphi^1_{rjr'pish} = 1 - \left[ (1 - \beta_r^{s,h,1} e_c)^{\chi_h} (1 - \beta_r^{s,h,1})^{\bar{\chi}_h} \right]$$

- To women/uncircumcised men on PrEP:

$$\varphi^2_{rjr'pish} = 1 - \left[ (1-\beta_r^{s,h,2} e_c)^{\chi_h} (1-\beta_r^{s,h,2})^{\bar{\chi_h}} \right]$$

- To circumcised men not on PrEP:

$$\varphi^3_{MjFpish} = 1 - \left[ (1-\beta_M^{s,h,3} e_c)^{\chi_h} (1-\beta_M^{s,h,3})^{\bar{\chi_h}} \right]$$

- To circumcised men on PrEP:

$$\varphi^4_{MjFpish} = 1 - \left[ (1-\beta_M^{s,h,4} e_c)^{\chi_h} (1-\beta_M^{s,h,4})^{\bar{\chi_h}} \right]$$

where

$\beta_r^{s,h,x}$ is the transmission probability corresponding to the partner's disease stage, ART use and susceptible's PrEP/circumcision status (Table S1)

$\chi_h$ ($\bar{\chi}_h$) denotes the proportion of coital acts (not) protected by condom use ($h = 1$ if no HCT/ART, $h = 2$ if HCT, $h = 3$ if ART), where, for $h = 1$ and $h = 3$,

$$\chi_h = f_i \eta_{jpi}$$

and

$$\bar{\chi}_h = (1 - f_i)\eta_{jpi}$$

In case the infected partner is aware of their status (i.e. assumed to reduce risk behaviour due to HIV counseling, $h = 2$),

$$\chi_2 = f_i \xi \eta_{jpi} \epsilon$$

and

$$\bar{\chi}_2 = (1 - f_i \xi)\eta_{jpi} \epsilon$$

where $\xi$ denotes the increase in condom use due to HCT, and $\epsilon$ denotes a decrease in the frequency of coital acts.

##### Intervention uptake rates

The rate of diagnosis $\tau^T$ is modelled such that the proportion of infected individuals that are aware of their status increases linearly over time until reaching a specified level $P_{HCT}$ in 2014. This is achieved as follows [6]:

- if $t < t_{start}$

$$\tau^T(t) = 0$$

- if $t_{start} < t \leq t_{start} + \frac{1}{r_T}$

$$\tau^T(t) = \left[(t - t_{start})r_T P_{HCT}\right] - \left[\frac{I^2(t) + I^3(t)}{I(t)}\right]$$

where $t_{start}$ is the time at which HIV testing is assumed to start (i.e. 1987), $1/r_T$ is the time it takes to reach the desired baseline coverage level $P_{HCT}$ in 2014, and $\left[\frac{I^2(t)+I^3(t)}{I(t)}\right]$ is the proportion aware of their HIV+ status at time $t$. In our baseline model (i.e. the model run for an additional 15 years without intervention scale-up), $P_{HCT}$ is slightly increased to 65% to avoid a zero rate of treatment uptake. In the model with intervention scale-up, the rate of diagnosis is increased from 2014 onward such that the proportion aware of their HIV+ status in 2029 reaches a specified level $P_{HCT2}$ as

- if $t > t_{start} + \frac{1}{r_T}$

$$\tau^T(t) = \tau^T(t-1) + \left[\left(t - (t_{start} + \frac{1}{r_T})\right)r_{T2}(P_{HCT_2} - P_{HCT})\right] - \left[\frac{I^2(t) + I^3(t)}{I(t)} - pHCT\right]$$

where $1/r_{T2}$ is the duration of scale-up.

A proportion of individuals, $P_{ART}$, aware of their HIV-positive status initiate ART on average one year after diagnosis, resulting in a monthly ART uptake rate of

$$\tau_A^s(t) = \left(1 + (-log(1 - P_{ART}(t)))\right)^{\frac{1}{12}} - 1$$

where $P_{ART}$ is assumed to increase over time according to a Hill function until reaching a specified baseline value.

$$P_{ART} = \begin{cases} 0 & \text{if } t < t_1 \\ P_{max}^{art} \frac{(t-t_1)^2}{(t-t_1)^2 + 10^2} & \text{if } t_1 \leq t \leq t_2 \\ P_{max}^{art} + (P_{max.up}^{art} - P_{max}^{art}) \frac{(t-t_2)^6}{(t-t_2)^6 + 95^6} & \text{if } t > t_2 \end{cases}$$

where $P_{max}^{art}$ is the desired probability of ART initiation at $t_2$ (i.e. 2014), and $P_{max.up}^{art}$ is the desired probability of ART initiation after scale-up (i.e. 2029). For infected individuals in the acute and chronic stages of infection $t_1 = 2011$, while for those in the (pre-)AIDS stage $t_1 = 2004$.

Before 2014, the rate of HCT uptake in susceptible individuals is assumed to be equal to the rate of diagnosis, hence the rate of VMMC uptake before 2014 and in our baseline scenario is

$$\sigma(t) = \left[\left(1 + (-log(1 - P_{CIRC}(t)))\right)^{\frac{1}{12}} - 1\right] HCT_m \tau^T(t)_{\{t \leq 2014\}}$$

where $HCT_m$ represents reduced HCT uptake in males and $P_{CIRC}$ is the proportion of individuals accepting VMMC on average one year after being tested. As such we assume there was no VMMC before the introduction of HIV testing, although a constant proportion $p_c$ of males enter the population already circumcised. The probability $P_{CIRC}$ of accepting VMMC increases over time according to a Hill function until reaching a specified value.

$$P_{CIRC} = \begin{cases} 0 & \text{if } t < t_1 \\ P^{circ}_{max} \frac{(t-t_1)^4}{(t-t_1)^4 + 100^4} & \text{if } t_1 \leq t \leq t_2 \\ P^{circ}_{max} + (P^{circ}_{max.up} - P^{circ}_{max}) \frac{(t-t_2)^6}{(t-t_2)^6 + 85^6} & \text{if } t > t_2 \end{cases}$$

where $P^{circ}_{max}$ is the desired probability of accepting VMMC at $t_2$ (i.e. 2014), and $P^{circ}_{max.up}$ is the desired probability of accepting VMMC after scale-up (i.e. 2029). In the models with intervention scale-up, HCT uptake in susceptible individuals is given by

$$\tau^{ST}(t) = P^S_{HCT} \frac{(t - t_1)^5}{(t - t_1)^5 + 100^5}$$

where $P^S_{HCT}$ is the desired proportion of susceptible individuals accepting HCT after intervention scale-up, and $t_1 = 2014$. The rate of VMMC uptake after 2014 then becomes

$$\sigma(t) = \left[\left(1 + (-log(1 - P_{CIRC}(t)))\right)^{\frac{1}{12}} - 1\right] HCT_m \tau^{ST}(t).$$

PrEP uptake is only included in the model with intervention scale-up and is modelled similarly, with $P_{PrEP}$ being the proportion of susceptible individuals initiating PrEP on average one year after testing, which increases over time until reaching a specified level $P^{prep}_{max}$,

$$P_{PrEP} = \begin{cases} 0 & \text{if } t < t_1 \\ P^{prep}_{max} \frac{(t-t_1)^4}{(t-t_1)^5 + 85^5} & \text{if } t \geq t_1 \end{cases}$$

where $t_1 = 2015$. The rate of PrEP uptake then becomes

$$\theta_M(t) = \left[\left(1 + (-log(1 - P_{PrEP}(t)))\right)^{\frac{1}{12}} - 1\right] HCT_m \tau^{ST}(t)$$

for males, and

$$\theta_F(t) = \left[\left(1 + (-log(1 - P_{PrEP}(t)))\right)^{\frac{1}{12}} - 1\right] \tau^{ST}(t)$$

for females.

Condom use is assumed to be different for each type of partnership and increases over time according to a Hill function

$$f_i(t) = \begin{cases} f_{i_{max}} \frac{(t-0)^4}{(t-0)^4+200^4} & \text{if } t < t_1 \\ f_{i_{max}} + (f_{i_{max.up}} - f_{i_{max}}) \frac{(t-t_1)^6}{(t-t_1)^6+85^6} & \text{if } t \geq t_1 \end{cases}$$

where $f_{i_{max}}$ is the maximum (baseline) level of condom use. After 2014, $f_{i_{max}}$ is increased to $f_{i_{max.up}}$ representing intervention scale-up.

##### Model calibration

The model was run for 45 years using a time step of one month for each of the 10 000 LHS-generated parameter input sets $\theta$ obtained from the uncertainty ranges in Table S2. Overall goodness of fit (GOF) was measured as the sum of the squared deviations from the target prevalence in each risk group, $p_{rj}$, and the total prevalence $p$,

$$GOF = (\hat{p} - p)^2 + \sum (\hat{p_{rj}} - p_{rj})^2$$

We used an active learning approach to reduce the uncertainty ranges of the model parameters in an iterative way. After each iteration, the subspace of parameters leading to the top 1% of model outputs (in terms of GOF), $\Theta_{1\%}$ was investigated using several methods and where possible the uncertainty ranges were reduced.

The activity region finder (ARF) proposed by Amaratunga and Cabrera was first used to investigate the subspace $\Theta_{1\%}$.[7] ARF is a recursive partitioning classification tree method where each of the 10 000 LHS-generated parameter sets are considered as input, and their corresponding binary indicators that equal 1 if $\theta \in \Theta_{1\%}$ and 0 otherwise are considered as output. ARF then shows which input parameter sets occur more frequently in the subspace $\Theta_{1\%}$ and thus provide a better fit to the observed prevalence data. The maximal information coefficient (MIC) was then used to quantify the association structure between the parameters in the subspace $\Theta_{1\%}$.[8] The MIC ranges between 0 and 1, with higher values indicating a stronger relationship between the parameters. In this way, insight was obtained on which parameters are highly correlated and thus would be more difficult to estimate. Additionally, influence of the data on the estimated parameters was identified by univariately exploring the subspace $\Theta_{1\%}$. For each parameter a graphical comparison of the (prior) density of the uniform distribution $f_U$ (i.e. the range as in Table S2) with the (posterior) density $f_{\Theta_{1\%}}$, corresponding to the values within the subspace $\Theta_{1\%}$, was made.[9] A peaked unimodal density $f_{\Theta_{1\%}}$ indicates that the parameter is (highly) informed by the data. In case a parameter's updated uncertainty range would be small and the parameter appeared to be not highly informed by the data, this parameter was fixed to its value in the current 'best' solution. This active learning process was repeated until an optimal value for the parameters was obtained or until the parameter ranges were stable in case they could not be reduced to a single value.

For most parameters, stability in uncertainty ranges was achieved after 24 iterations, and the best GOF (lowest discrepancy between observed and simulated prevalence) was obtained. Model parameters corresponding to the 'best fit' model are shown in Table S2. In the final iteration, the highest MIC was 0.56, measuring the association between $\eta_{33main}$ (frequency of main partnership coital acts among individuals from the high risk group) and $x_{casual}$ (constant controlling the extent to which each gender compromises with the wishes of the other gender, for casual partnerships). MIC was also high between $\beta_m$ and $P_{HCT}$ (0.50) and between $f_{casual}$ and $\eta_{33main}$ (0.46). All other MIC were smaller than 0.40, indicating no substantial association between the remaining parameters in the subspace $\Theta_{1\%}$. The unimodal peaks in Figure S2 show that some of these parameters with high MIC also appear to be quite informed by the data. In addition, these parameters also often took part in the ARF process, further indicating the difficulty in reducing their ranges. Figure S3 shows the evolution of HIV prevalence (a) and incidence (b) under the 'best fit' model without increasing intervention uptake after 2014.

**Assessing the impact of intervention scale-up**

As baseline, we run the calibrated model for an additional 15 years (2014-2029) without intervention scale-up (i.e. assuming uptake remains constant from 2014 onward). To assess the impact of different combinations of increasing intervention uptake, we use LHS to sample 500 input parameter sets $\theta_I$ from the ranges of the intervention parameters (Table S3). We first investigate the impact of intervention scale-up by running the 'best fit' model for an additional 15 years (2014-2029). Different combinations of intervention scale-up were compared in terms of their impact on the relative reduction in HIV incidence and prevalence, AIDS-related mortality, and the total number of new infections, by comparing the model with intervention scale-up to the baseline model without increased intervention uptake. To account for the remaining uncertainty in the baseline model parameters, parameter ranges of simulated epidemics belonging to the top 1% of model outputs, $\Theta_{1\%}$, were also used to investigate the impact of combined interventions.

Intervention impact was evaluated on the following model output (as % reduction), per subgroup $(r, j)$ and overall.

Prevalence: $P_{rj}(t) = \frac{\sum_{h=1}^{3}(H_{rj}^h(t)+Y_{rj}^h(t)+Z_{rj}^h(t)+A_{rj}^h(t))}{N_{rj}}$

New infections: $NI_{rj}(t) = \sum_x \pi_{rj}^x(t) S_{rj}^x(t)$

Incidence (per 100 PY): $IR(t) = \frac{\sum_{x,r,j} \pi_{rj}^x S_{rj}^x}{\sum_{x,r,j} S_{rj}^x} \times 100 \times 12$

AIDS-related mortality (per 100 PY): $\Delta_{rj}(t) = \frac{\sum_{h=1}^{3}(\delta^h A_{rj}^h(t))}{N_{rj}(t)} \times 100 \times 12$

**Table S1. Probability of transmission per coital act, where baseline corresponds to the chronic LV stage.**

| Disease stage | ART | ART+PrEP | ART+CIRC | ART+PrEP+CIRC | PrEP | CIRC | CIRC+PrEP |
|---|---|---|---|---|---|---|---|
| Baseline: $\beta_r^{1,1,1}$ | $\beta_r^{1,3,1}$ | $\beta_r^{1,3,2}$ | $\beta_M^{1,3,3}$ | $\beta_M^{1,3,4}$ | $\beta_r^{1,1,2}$ | $\beta_M^{1,1,3}$ | $\beta_M^{1,1,4}$ |
| Initial HV: $\beta_r^{2,1,1}$ | $\beta_r^{2,3,1}$ | $\beta_r^{2,3,2}$ | $\beta_M^{2,3,3}$ | $\beta_M^{2,3,4}$ | $\beta_r^{2,1,2}$ | $\beta_M^{2,1,3}$ | $\beta_M^{2,1,4}$ |
| pre-AIDS LV: $\beta_r^{3,1,1}$ | $\beta_r^{3,3,1}$ | $\beta_r^{3,3,2}$ | $\beta_M^{3,3,3}$ | $\beta_M^{3,3,4}$ | $\beta_r^{3,1,2}$ | $\beta_M^{3,1,3}$ | $\beta_M^{3,1,4}$ |
| AIDS HV: $\beta_r^{4,1,1}$ | $\beta_r^{4,3,1}$ | $\beta_r^{4,3,2}$ | $\beta_M^{4,3,3}$ | $\beta_M^{4,3,4}$ | $\beta_r^{4,1,2}$ | $\beta_M^{4,1,3}$ | $\beta_M^{4,1,4}$ |

#### Table S2. Uncertainty ranges and fixed values for the baseline model parameters and value in the 'best fit' solution.

| Parameter | Range / Value | | Reference | Best fit | |
|---|---|---|---|---|---|
| | Male | Female | | Male | Female |
| **Demographic** | | | | | |
| Initial population size (N) | 2000 | | | | |
| Population size per class | | | | | |
| low risk | 0.33*N | 0.43*N | HIVCOMB 2014 census | | |
| medium risk | 0.07*N | 0.02*N | HIVCOMB 2014 census | | |
| high risk | 0.10*N | 0.05*N | HIVCOMB 2014 census | | |
| Mortality/recruitment rate ($\psi$) | 0.0019 | | Based on 45 years of sexual activity | | |
| Population growth rate ($\mu$) | 0.0021 | | Based on yearly growth rate of 2.5% | | |
| **Epidemiologic** | | | | | |
| Initial HIV prevalence | 1% | | | | |
| Untreated disease progression | | | | | |
| Average duration of initial HV stage ($1/\nu^1$) | 3.25 months | | [10] | | |
| Average duration of LV stage ($1/\Omega^1$) | 100 months | | | | |
| Average duration of pre-AIDS stage ($1/\Psi^1$) | 14 months | | | | |
| Average duration of AIDS stage ($1/\delta^1$) | 16 months | | | | |
| Treated disease progression | | | | | |
| Average duration of initial HV stage ($1/\nu^2$) | 98 months | | [10] | | |
| Average duration of LV stage ($1/\Omega^2$) | 648 months | | | | |
| Average duration of pre-AIDS stage ($1/\Psi^2$) | 30 months | | | | |
| Average duration of AIDS stage ($1/\delta^2$) | 33 months | | | | |
| **Transmission** | | | | | |
| Transmission probability per sex act ($\beta_r$) | $0.001 - 0.003$ | | [11] | 0.001 | 0.002 |
| Multiplication factor (treated = untreated) | | | | | |
| initial HV ($\alpha_1$) | 26 | | [12] | | |
| pre-aids HV ($\alpha_2$) | 7 | | [12] | | |
| AIDS stage ($\alpha_3$) | 1 | | | | |
| **Intervention** | | | | | |
| Efficacy of ART ($e_a$) | 0.90 | | Reduction in infectiousness (Li et al) | | |
| Efficacy of PrEP ($e_p$) | 0.67 | | 67% reduction in susc. [13] | | |
| PrEP adherence ($P_r$) | 0.8 | | Assumed | | |
| Efficacy of condom use ($e_c$) | 0.1 | | Li et al (relative infectiousness) | | |
| Efficacy of circumcision ($e_{circ}$) | 0.60 | | Reduction in susceptibility [14] | | |
| Rate of PrEP uptake ($\theta$) | 0 | | No PrEP in baseline model | | |
| Rate of PrEP discontinuation ($\omega$) | 0 | | - | | |
| Maximum probability of accepting ART ($P^{art}_{max}$) | | | | | |
| Acute stage | 0.05 - 0.3 | | Assumed | 0.157 | |
| Chronic stage | 0.05 - 0.3 | | Assumed | 0.192 | |
| Pre-AIDS stage | 0.1 - 0.3 | | Assumed | 0.185 | |
| AIDS stage | 0.2 - 0.6 | | Twice as fast as in pre-AIDS stage | 0.371 | |
| Rate of ART dropout ($\gamma$) | 0.0186 | | Annual dropout of 20% [1] | | |
| Proportion accepting VMMC ($P^{circ}_{max}$) | - | 0.1 - 0.5 | Assumed annual proportion of 10-50% | 0.313 | |
| Proportion already circumcised ($p_c$) | - | 0.1 - 0.2 | [15] | 0.318 | |
| Proportion aware of HIV+ status in 2014 ($P_{HCT}$) | 0.5 - 0.7 | | Assumed | 0.634 | |
| Relative HCT uptake in males | 0.5 - 0.9 | - | Assumed based on UPHIA report 2016 | 0.700 | |
| Relative ART uptake in males | 0.5 - 0.95 | - | Assumed based on UPHIA report 2016 | 0.759 | |
| Relative number of sex acts due to HCT ($\epsilon$) | 0.9 | | Assume 10% reduction | | |
| Increase in condom use due to HCT ($\xi$) | 1.10 | | Assume 10% increase | | |
| Relative number of sex acts due to untreated AIDS ($\epsilon_{AIDS}$) | 0.1 | | Assumed | | |
| **Behavioral** | | | | | |
| Probability of condom use per sex act ($f_{i_{max}}$) | | | | | |
| Main partnerships | 0.01 - 0.1 | | Assumed | 0.070 | |
| Non-main regular partnerships | 0.1 - 0.3 | | Assumed | 0.261 | |
| Casual partnerships | 0.3 - 0.6 | | HIVCOMB 2014 census | 0.428 | |
| Number of casual sex acts per month ($\eta_{rj_{casual}}$) | | | | | |
| high-high risk | 1-5 | | Assumed | 3.586 | |
| Number of non-main regular sex acts per month ($\eta_{rj_{regular}}$) | | | | | |
| medium-medium risk | 1-5 | | Assumed | 4.338 | |
| medium-high risk | 1-5 | | Assumed | 5.535 | |
| high-medium risk | 1-5 | | Assumed | 8.429 | |
| high-high risk | 1-5 | | Assumed | 2.587 | |
| Number of main sex acts per month ($\eta_{rj_{main}}$) | | | | | |

| | | | | |
|---|---|---|---|---|
| low-low risk | 1-30 | Assumed | 19.843 | |
| low-medium risk | 1-30 | Assumed | 16.364 | |
| low-high risk | 1-30 | Assumed | 8.925 | |
| medium-low risk | 1-20 | Assumed | 6.471 | |
| medium-medium risk | 1-20 | Assumed | 4.834 | |
| medium-high risk | 1-20 | Assumed | 5.835 | |
| high-low risk | 1-15 | Assumed | 5.676 | |
| high-medium risk | 1-15 | Assumed | 4.808 | |
| high-high risk | 1-15 | Assumed | 5.469 | |
| Rate of partner acquisition for main partnerships ($d_{rj_{main}}$) | | | | |
| low risk | 0.0083 - 0.4167 | Assuming 5 new main partners per year to one every 10 years | 0.114 | 0.347 |
| medium risk | 0.0083 - 0.4167 | Assuming 5 new main partners per year to one every 10 years | 0.339 | 0.266 |
| high risk | 0.0083 - 0.4167 | Assuming 5 new main partners per year to one every 10 years | 0.187 | 0.225 |
| Rate of partner change for regular partnerships ($d_{rj_{regular}}$) | | | | |
| medium risk | 0.4167 - 0.8333 | Assuming 5-10 new partners every year | 0.708 | 0.604 |
| high risk | 0.4167 - 0.8333 | Assuming 5-10 new partners every year | 0.547 | 0.571 |
| Rate of partner change for casual partnerships ($d_{rj_{casual}}$) | | | | |
| high risk | 0.6667 - 1 | Assuming 8-12 new partners every year | 0.944 | 0.766 |
| Degree of assortativity ($\mathcal{A}_i$) | | | | |
| Main partnerships | $0.5 - 0.9$ | Assumed tendency towards assortative mixing | 0.573 | |
| Non-main regular partnerships | $0.1 - 0.5$ | Assumed tendency towards random mixing | 0.216 | |
| Casual partnerships | 1 | Motivated in text | | |
| Constant to adjust partnership imabalance ($x_i$) | | | | |
| Main partnerships | 0.2 - 0.8 | Assumed | 0.301 | |
| Non-main regular partnerships | 0.2 - 0.8 | Assumed | 0.394 | |
| Casual partnerships | 0.2 - 0.8 | Assumed | 0.201 | |

**Table S3. Uptake ranges for the intervention parameters and their values in the highest impact scenario.**

| Intervention | Range / Value Male | Female | Reference | Highest impact scenario | Baseline model |
|---|---|---|---|---|---|
| Efficacy of PrEP ($e_p$) | 0.67 | | 67% reduction in susc. [13] | | |
| PrEP adherence ($P_r$) | 0.6 | | Assumed | | |
| Probability of initiating PrEP ($P_{max}^{prep}$) | 0.2 - 0.8 | | Assumed | 0.3040 | - |
| Proportion of susceptibles accepting HCT ($P_{HCT}^{S}$) | 0.2 - 0.8 | | Assumed | 0.4615 | - |
| Rate of PrEP discontinuation ($\omega$) | 0.0186 | | Annual dropout of 20% | | |
| Probability of initiating ART ($P_{max.up}^{art}$) | | | | | |
| Acute stage | 0.2 - 0.8 | | Assumed | 0.3315 | 0.157 |
| Chronic stage | 0.2 - 0.8 | | Assumed | 0.7251 | 0.192 |
| Pre-AIDS stage | 0.2 - 0.8 | | Assumed | 0.3985 | 0.185 |
| AIDS stage | 0.4 - 0.8 | | Assumed | 0.5197 | 0.371 |
| Rate of ART dropout ($\gamma$) | 0.0186 | | Annual dropout of 20% [1] | | |
| Proportion accepting VMMC ($P_{max.up}^{circ}$) | - | 0.3 - 0.8 | Assumed | 0.6305 | 0.313 |
| Proportion aware of HIV+ status in 2029 ($P_{HCT_2}$) | 0.6 - 0.9 | | Assumed | 0.8944 | - |
| Probability of condom use per sex act ($f_{i_{max.up}}$) | | | | | |
| Main partnerships | 0.1 - 0.6 | | Assumed | 0.5806 | 0.070 |
| Non-main regular partnerships | 0.3 - 0.6 | | Assumed | 0.5818 | 0.261 |
| Casual partnerships | 0.5 - 0.6 | | Assumed | 0.585 | 0.428 |

#### Table S4. Sensitivity analyses for the impact of combined intervention.

| Scenario | Relative reduction compared to baseline | | | |
|---|---|---|---|---|
| | Incidence | Prevalence | AIDS-related mortality | New infections over 15 years |
| PrEP adherence of 90% | 0.8680 | 0.3815 | 0.5378 | 0.3987 |
| PrEP adherence of 30% | 0.8657 | 0.3805 | 0.5374 | 0.4119 |
| PrEP dropout rate of 5% | 0.8676 | 0.3812 | 0.5377 | 0.4125 |
| No PrEP use | 0.8651 | 0.3802 | 0.5373 | 0.3976 |
| No increase in condom use | 0.5619 | 0.1559 | 0.4111 | 0.2063 |
| ART dropout rate of 5% from 2014 onward | 0.9116 | 0.3915 | 0.6368 | 0.4751 |
| ART dropout rate of 70% from 2014 onward | 0.7778 | 0.3637 | 0.3407 | 0.2999 |
| Equal uptake in males and females | 0.8811 | 0.3980 | 0.5647 | 0.4421 |
| Probability of 30% of ART initiation in chronic stage | 0.8154 | 0.3783 | 0.4154 | 0.3805 |

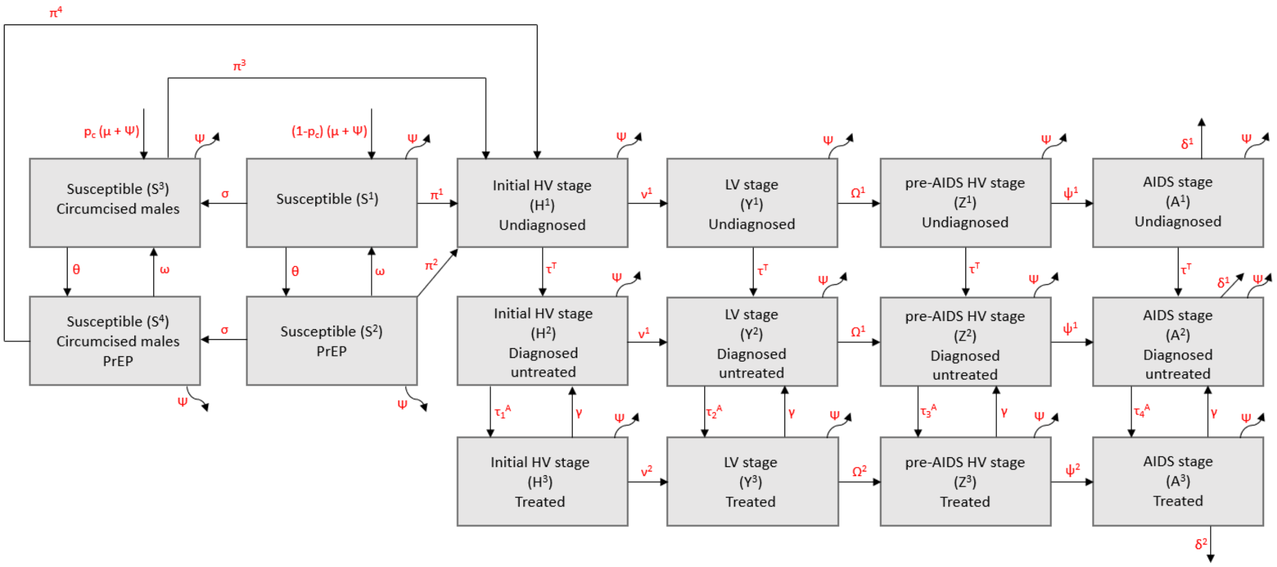


Figure S1: Model structure for males in risk group $j$ (which is the same for females, without circumcision compartments)

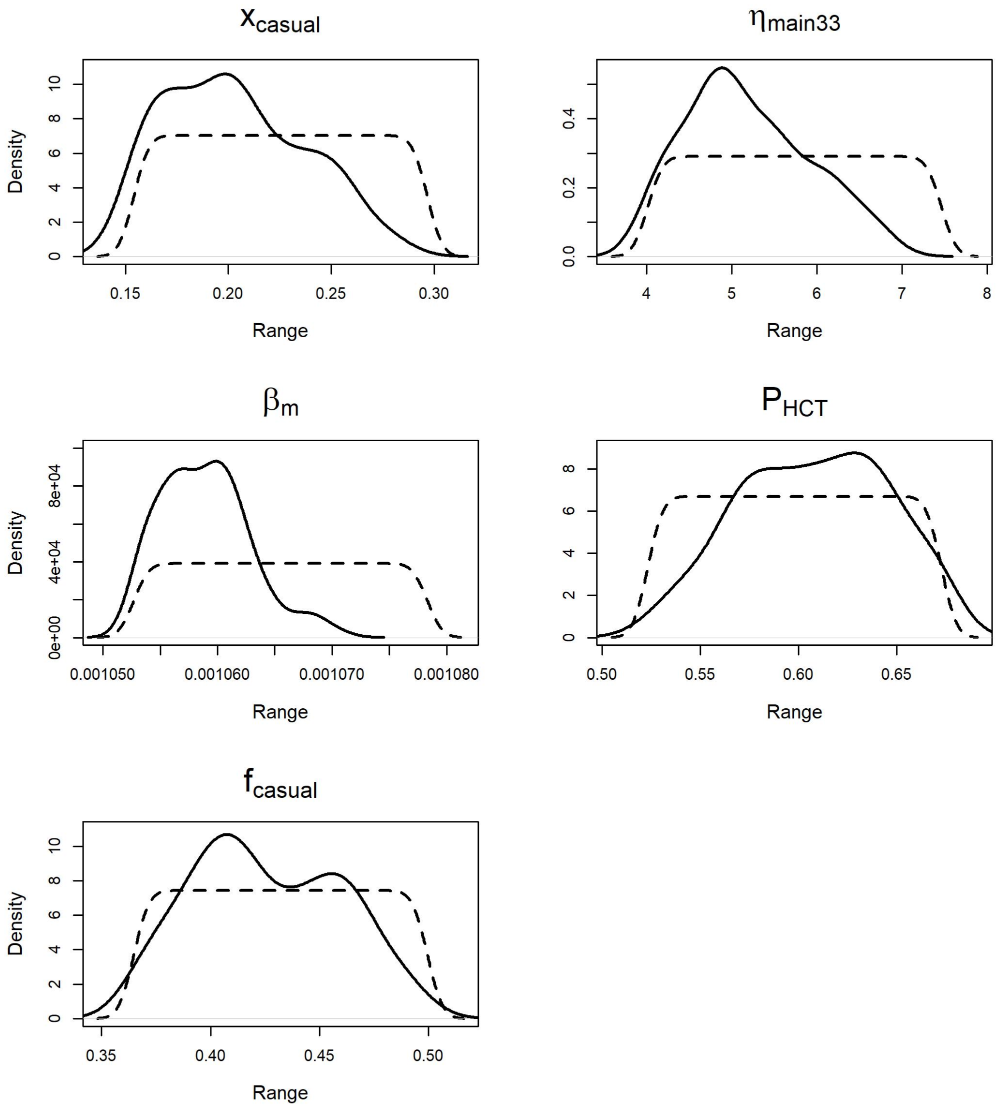


Figure S2: Parameters that are highly informed by the data. Dashed lines represent the density $f_U$ of the uniform distribution on the uncertainty range of the input parameter. Solid lines represent the density $f_{\Theta 1\%}$ corresponding to the values within the subspace $\Theta_{1\%}$. A peaked unimodal density $f_{\Theta 1\%}$ indicates that the parameter was highly informed by the data.

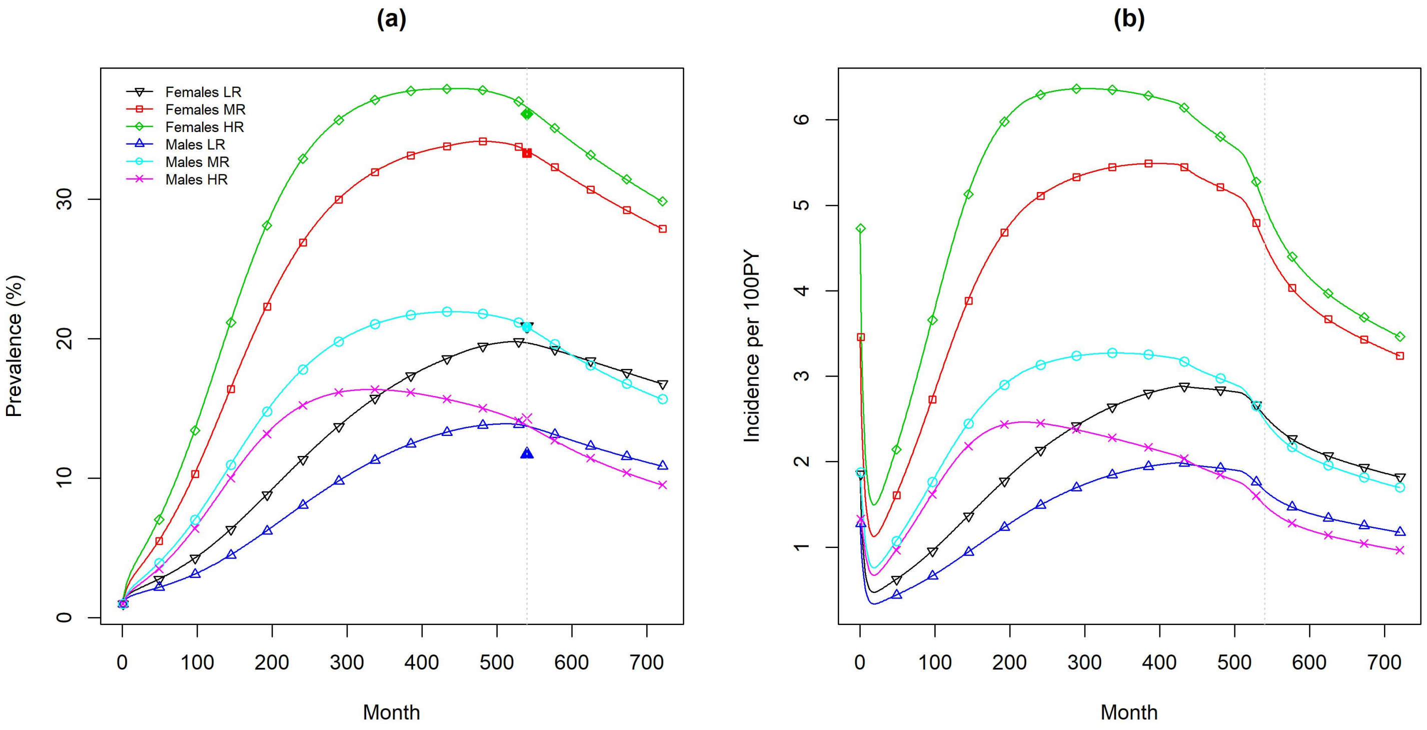


Figure S3: HIV prevalence (a) and incidence (b) in the different sexual risk groups obtained from the 'best fit' model. Filled symbols on the dotted grey vertical line represent the observed prevalence in 2014 obtained from the HIVCOMB study, to which the model was calibrated.

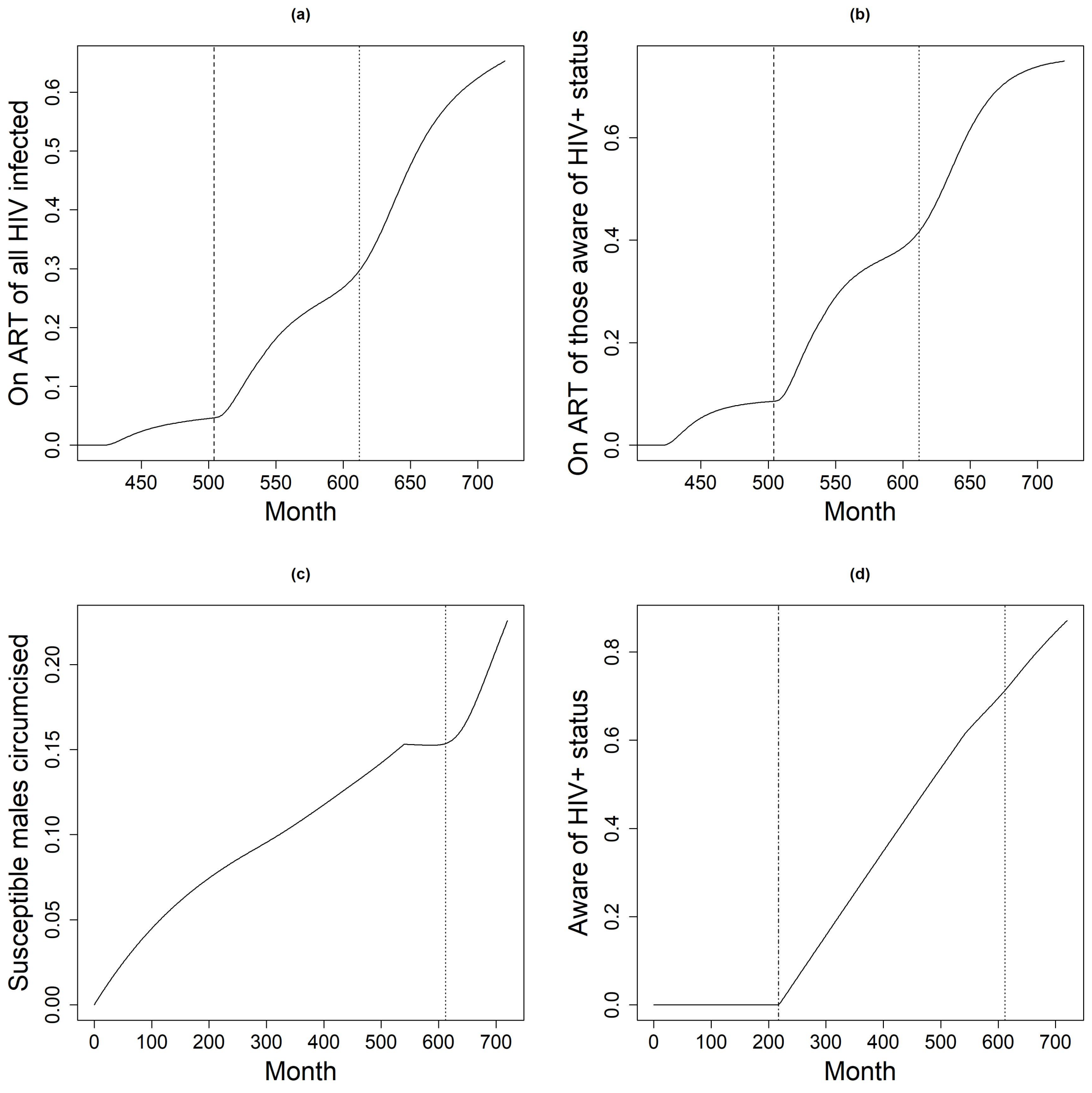


Figure S4: Intervention coverage over time. (a) HIV infected individuals on ART, (b) HIV infected individuals aware of their status on ART, (c) circumcised susceptible males, and (d) individuals aware of their HIV+ status. Dashed lines indicate the year 2011 when ART was implemented in the chronic stage, dotted lines indicate the year 2020 when intervention scale-up was started, dash-dotted lines indicate 1987 when HIV testing was introduced.

[11] M. C. Boily, R. F. Baggaley, L. Wang, B. Masse, R. G. White, R. Hayes, and M. Alary. Heterosexual risk of HIV-1 infection per sexual act: a systematic review

and meta-analysis of observational studies. *Lancet Infectious Diseases*, 9:118–129, 2009.

[12] T. D. Hollingsworth, R. M. Anderson, and C. Fraser. HIV-1 transmission, by stage of infection. *Journal of Infectious Diseases*, 198:687–693, 2008.

[13] J. M. Baeten, D. Donnell, and P. Ndase et al. Antiretroviral prophylaxis for HIV prevention in heterosexual men and women. *The New England Journal of Medicine*, 367:399–410, 2012.

[14] R. H. Gray, G. Kigozi, and D. Serwadda et al. Male circumcision for HIV prevention in men in Rakai, Uganda: a randomized trial. *Lancet*, 369:657–666, 2007.

[15] Uganda Bureau of Statistics. The national population and housing census 2014 - main report. 2016.
